# Protein haploinsufficiency drivers identify *MYBPC3* mutations that cause hypertrophic cardiomyopathy

**DOI:** 10.1101/2020.05.04.20087726

**Authors:** Carmen Suay-Corredera, Maria Rosaria Pricolo, Elías Herrero-Galán, Diana Velázquez-Carreras, David Sánchez-Ortiz, Diego García-Giustiniani, Javier Delgado, Juan José Galano-Frutos, Helena García-Cebollada, Silvia Vilches, Fernando Domínguez, María Sabater Molina, Roberto Barriales-Villa, Giulia Frisso, Javier Sancho, Luis Serrano, Pablo García-Pavía, Lorenzo Monserrat, Jorge Alegre-Cebollada

**Author notes:** These authors contributed equally to this work (Carmen Suay-Corredera, Maria Rosaria Pricolo). To whom correspondence should be addressed (@AlegreCebollada).

## Abstract

Hypertrophic cardiomyopathy (HCM) is the most common inherited cardiac disease. Mutations in *MYBPC3*, the gene encoding cardiac myosin-binding protein C (cMyBP-C), are a leading cause of HCM. However, it remains challenging to define whether specific gene variants found in patients are pathogenic or not, limiting the reach of cardiovascular genetics in the management of HCM. Here, we have examined cMyBP-C haploinsufficiency drivers in 68 clinically annotated non-truncating variants of *MYBPC3*. We find that 45% of the pathogenic variants show alterations in RNA splicing or protein stability, which can be linked to pathogenicity with 100% and 94% specificity, respectively. Relevant for variant annotation, we uncover that 9% of non-truncating variants of *MYBPC3* currently classified as of uncertain significance induce one of these molecular phenotypes. We propose that alteration of RNA splicing or protein stability caused by *MYBPC3* variants provide strong evidence of their pathogenicity, leading to improved clinical management of HCM patients and their families.

## INTRODUCTION

Hypertrophic cardiomyopathy (HCM) is the most frequent inherited cardiac muscle disease, with an estimated global prevalence of at least 0.5% ^1–3^. HCM is a frequent cause of sudden cardiac death in the young, and can result in several cardiovascular complications including heart failure and thromboembolism ^1^. Identification of HCM-causative mutations has revolutionized clinical management of patients and their families. Nowadays, genetic testing can confirm a clinical suspicion, help differential diagnosis, and is at the basis of family cascade screening allowing reproductive and professional counselling. However, similar to many other genetic conditions, a fundamental limitation stems from the rarity of HCM genetic variants^4,5^. For many of them, the number of individuals who can be studied is scarce. As a result, detailed segregation studies are seldom possible, leaving numerous variants classified as of uncertain significance (VUS). This situation has motivated efforts to assess pathogenicity of VUS using functional and population genetics approaches ^6–10^.

Mutations in *MYBPC3*, the gene coding for cardiac myosin-binding protein C (cMyBP-C), are a leading cause of HCM (**Figure 1a**) ^4,11,12^. Most well-established pathogenic variants in *MYBPC3* are frameshift, nonsense or conserved RNA splice site mutations that result in truncated polypeptides, which are more prone to degradation leading to lower total cMyBP-C protein levels (haploinsufficiency) ^13,14^ Indeed, cMyBP-C haploinsufficiency results by itself in the development of HCM (**Figure 1a**, ***middle***) ^14–17^. Intriguingly, up to 15% of familial HCM cases can be caused by mutations in exonic regions of *MYBPC3* that do not lead to truncations (**Figure 1a**, ***right***) ^3,4,14,16,18^, including the most common pathogenic mutation in HCM (c.1504C>T, p.R502W) ^13^. As of April 2020, ClinVar lists 708 missense and synonymous variants in *MYPBC3* potentially linked to HCM.

**Figure 1.**
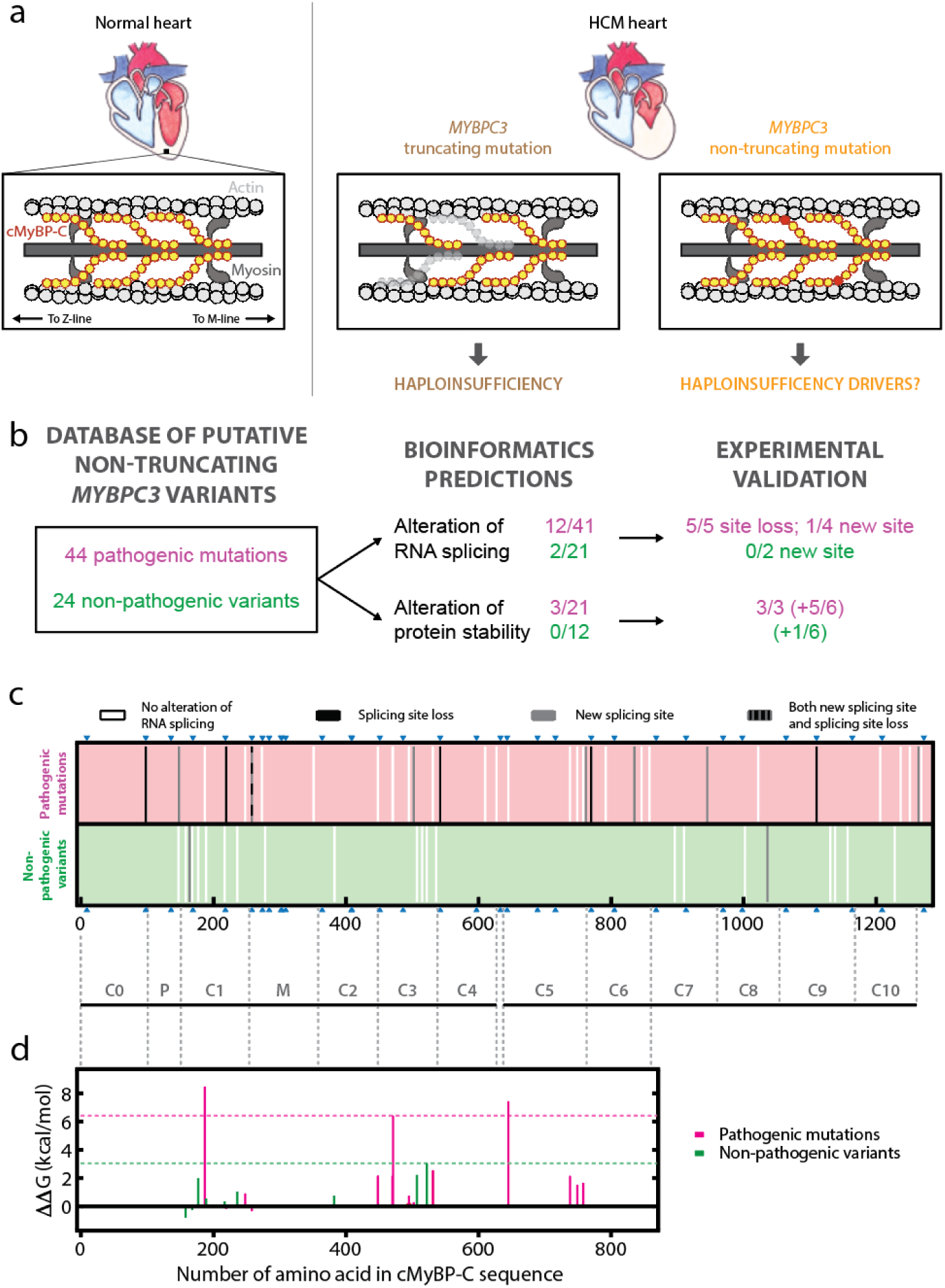
cMyBP-C haploinsufficiency drivers induced by pathogenic and non-pathogenic variants of *MYBPC3*. **(a)** *Left*: scheme of the location of cMyBP-C (in yellow) in the sarcomere. *Middle*: Most HCM-causing mutations in *MYBPC3* lead to truncated polypeptides and protein haploinsufficiency. *Right:* The remaining mutations are non-truncating and result in full-length mutant proteins (mutated domain is represented in red). **(b)** Workflow to identify cMyBP-C haploinsufficiency drivers in pathogenic and non-pathogenic variants of *MYBPC3*. The number of variants that are positive for predicted alterations of RNA splicing or protein stability are indicated, together with the outcomes of experimental validation (the protein stability of 6 variants per group were also screened, shown between parentheses). Pathogenic mutations are indicated in pink and non-pathogenic variants, in green. **(c)** Prediction of alterations in RNA splicing. Each bar corresponds to a single variant and is colored according to the predicted effect. Predictions for pathogenic mutations are shown in the upper half of the panel, while results for non-pathogenic variants appear in the lower half. Blue triangles indicate exon-exon boundaries. Organization of domains in cMyBP-C is indicated at the bottom of the panel. **(d)** Prediction of alterations of protein stability. Each bar corresponds to a single variant and is colored according to the pathogenicity of the variant (pathogenic, pink; non-pathogenic, green). The dotted lines mark the highest destabilizing change in ΔΔG detected for a non-pathogenic variant (green), and the lowest destabilization of a pathogenic mutation over the non-pathogenic threshold (pink).

Non-truncating *MYBPC3* variants currently pose a major challenge to genetic diagnosis in HCM. Despite recent encouraging developments, for most variants it remains impossible to assign pathogenicity just from their location in the gene or the nature of specific amino acid changes ^10^. Interestingly, both truncating and non-truncating mutations lead to similar HCM clinical manifestation ^19,20^, suggesting that pathogenicity in non-truncating mutations could also result from cMyBP-C haploinsufficiency. Two mechanisms emerge as the most common inducers of protein haploinsufficiency by putative non-truncating variants in monogenic diseases, i.e. defects in RNA splicing that result in the appearance of premature stop codons^21^, and reduction of protein stability ^22^. Both haploinsufficiency drivers have been reported before in *MYBPC3* variants linked to HCM^7,8,16,18,23–26^. However, the association of these molecular features to disease remains unknown due to the absence of systematic comparison with non-pathogenic variants. For instance, mild changes in splicing may be better tolerated than full native splicing abrogation by a conserved splice site mutation^18^. Ito *et al*. have shown that putative non-truncating *MYBPC3* variants appearing in cardiomyopathy gene databases lead to more frequent RNA splicing alteration than variants in control databases, although interpretation of results is complicated by the presence of HCM mutation carriers in the general population ^7,27^.

Here, we have examined cMyBP-C haploinsufficiency drivers in missense and synonymous variants in *MYBPC3* in the quest of molecular features that can identify pathogenicity. Our strategy builds on robust pathogenicity assignment, which limits the number of variants that can be analyzed, but has the potential to identify and quantify pathogenic molecular phenotypes with high confidence. We propose that easily testable, mutation-induced RNA splicing alteration and extensive protein destabilization provide strong evidence of pathogenicity of close to 10% of putative non-truncating *MYBPC3* variants currently considered as of uncertain significance.

## RESULTS

### A database of clinically curated variants of *MYBPC3*

We built a database containing 44 pathogenic and 24 non-pathogenic putative synonymous and missense *MYBPC3* variants covering the entire coding sequence of the gene (see Methods, **Supplementary File S1**). These variants were selected because they have unambiguous pathogenicity assignment based only on cosegregation and epidemiological criteria validated by the American College of Medical Genetics (ACMG) ^28^. We verified that clinical information publicly available in ClinVar is in excellent agreement with our pathogenicity assignment (**Supplementary Figure S1**). In addition, all the non-pathogenic variants have minor allele frequency (MAF) >10^−4^, while that of the pathogenic variants is <10^−4^. This threshold has been proposed before on the basis of HCM prevalence in the general population ^13^. After configuration of the database of variants, we investigated whether alteration of RNA splicing and protein stability are specific to pathogenic mutations, under the premise that molecular features related to HCM should not appear in non-pathogenic variants. We first triaged variants using bioinformatics predictors and then validated positive hits experimentally (**Figures 1b-d,2,3**).

### Alteration of RNA splicing by pathogenic mutations

RNA splicing is the process by which non-coding introns are removed from precursor mRNA, leading to exon-only-containing mature mRNA. Splicing involves recognition of specific sequences, particularly the strictly conserved two first (donor site) and last (acceptor site) nucleotides of introns. Mutations in these sequences impair canonical splicing, resulting in alternative mRNAs that contain premature stop codons, or lead to insertion/deletions in the polypeptide ^29^. Indeed, mutations in *MYBPC3* that affect these conserved slicing sites are usually considered to lead to a null allele and are classified as pathogenic ^13^. However, the correct functioning of the splicing machinery also involves recognition of sequence features in the exons, particularly in regions close to intron-exon boundaries. As a consequence, mutations in exonic sequences of *MYBPC3* traditionally classified as missense can result in alterations of RNA splicing that lead to truncated polypeptides ^7,8,18,30^. Indeed, we find that 6 pathogenic mutations in our database are predicted *in silico* to induce splicing site losses (**Figure 1c, Table 1**). The 6 mutations target the first or last nucleotide of an exon (**Supplementary Note S1**). Predictions also identify 7 pathogenic mutations that can lead to activation of new splicing sites. In contrast, all non-pathogenic variants target nucleotides outside exon-exon boundaries and none of them is predicted to cause loss of native splicing sites. For two non-pathogenic variants, the appearance of a new splicing site is suggested (**Figure 1c, Table 1, Supplementary Note S1**).

**Table 1.** Experimental validation of predicted alterations of RNA splicing in pathogenic and non-pathogenic variants of *MYBPC3*. DL: donor loss; DG: donor gain; AL: acceptor loss; AG: acceptor gain. Validation of predictions using myocardial, blood or mini-gene experiments samples are indicated. Mutants c.292G>C, c.2503C>T, c.2834G>A and c.3790T>C could not be studied.

| Variant (cDNA) | Variant (Protein) | Pathogenic variant | Predicted splicing alteration | Experimental splicing alteration (myocardium) | Experimental splicing alteration (blood) | Experimental splicing alteration (mini-gene) | Reference |
| --- | --- | --- | --- | --- | --- | --- | --- |
| 292G>C | E98Q | Yes | DL | - | - | - | Supp. File S2 |
| 442G>A | G148R | Yes | AG | - | - | Yes | <sup>7</sup> |
| 492C>T | G164G | No | DG | - | - | No | Figure 2i |
| 655G>T | V219F | Yes | AL | - | - | Yes | <sup>7,36</sup> |
| 772G>A | E258K | Yes | DL, AG | Yes (DL) | Yes (DL) | Yes (DL) | <sup>7,8,18,37</sup> |
| 1505G>A | R502Q | Yes | AG | - | No | No | Figure 2b and <sup>7</sup> |
| 1624G>C | E542Q | Yes | DL | Yes | Yes | Yes | Figure 2b and <sup>7,16,18,30,36</sup> |
| 2286C>G | V762V | Yes | DG | - | - | No | Supp. Figure S2d |
| 2308G>A | D770N | Yes | DL | Yes | - | - | <sup>18</sup> |
| 2503C>T | R835C | Yes | DG | - | - | - | Supp. File S2 |
| 2834G>A | R945Q | Yes | AG | - | - | - | Supp. File S2 |
| 3106C>T | R1036C | No | AG | - | - | No | <sup>7</sup> |
| 3330G>A | M1110I | Yes | DL | - | Yes | - | Supp. Figure S2e |
| 3790T>C | C1264R | Yes | DG | - | - | - | Supp. File S2 |

To validate predicted alterations in RNA splicing, we mined data available in the literature and analyzed RNA splicing in those cases for which there was no reported experimental information. RNA splicing of the *MYBPC3* transcript is best studied from myocardial samples; however, the scarcity of genotyped myocardium has limited analysis to a few variants ^8,16,18^. We have employed two alternative, more accessible strategies whose results are so far in excellent agreement with experiments using myocardial samples, i.e. analysis of mRNA from the leukocyte fraction of peripheral blood from variant carriers ^30–35^, and *in vitro* experiments using mini-gene constructs ^7,34,36^ (**Supplementary Table S1**). In both cases, isolated mRNA was amplified by RT-PCR using specific pairs of primers, and results were analyzed by electrophoresis and Sanger sequencing (**Figure 2, Supplementary File S2**).

**Figure 2.**
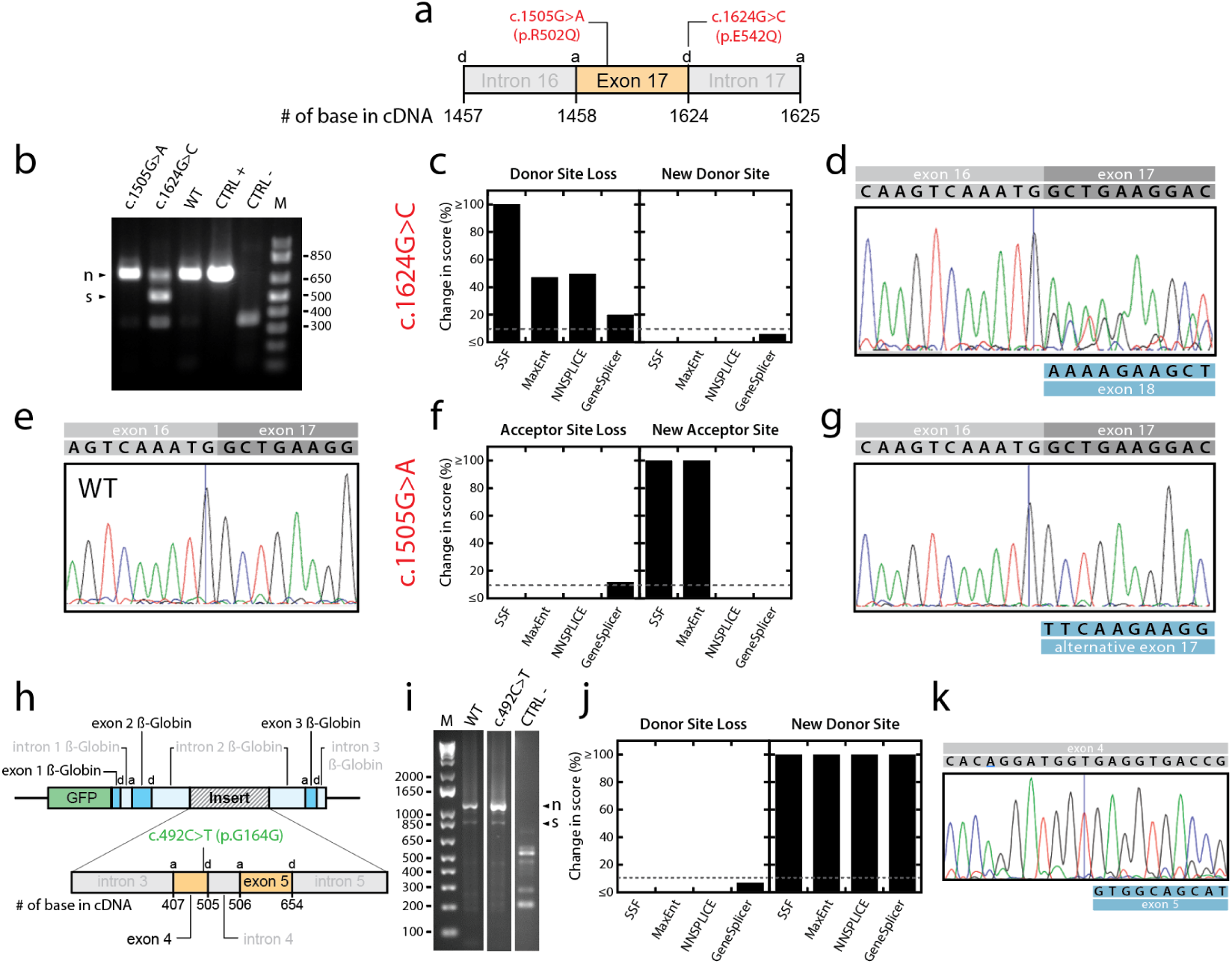
Experimental characterization of RNA splicing alteration induced by *MYBPC3* variants. **(a)** Location of two pathogenic mutations in the exon 17 of *MYBPC3* that are predicted to induce alterations of splicing. The positions of donor, d, and acceptor, *a*, splicing sites are indicated. **(b)** Experimental determination of RNA splicing by RT-PCR analysis of mRNA isolated from peripheral blood of carriers. CTRL+, mRNA obtained from healthy myocardium. CRTL-, mRNA isolated from HeLa cells, which do not express *MYBPC3*. The theoretical size of the amplified region if splicing is correct is 692 bp. In some samples, including CRTL –, a non-specific band is detected at a mobility between 300-400 bp. We could not identify the origin of this band. **(c)** Prediction of RNA splicing alteration for mutant c.1624G>C (p.E542Q). **(d)** Sanger sequencing result for c.1624G>C (p.E542Q) amplification product, which identifies the skipping of exon 17. The sequence resulting from the predicted change in splicing due to mutation c.1624G>C is shown in the blue box. Please note that the sequences of exons 17 and 18 are detected from the last nucleotide of exon 16, as expected from the presence of wild-type and mutated alleles in the heterozygous donor. **(e)** Sanger sequencing result for the WT amplification product showing normal splicing. **(f)** Prediction of RNA splicing alteration for mutant c.1505G>A (p.R502Q). **(g)** Sanger sequencing result for c.1505G>A (p.R502Q). **(h)** Mini-gene strategy to study splicing defects in non-pathogenic variant c.492C>T (p.G164G), which is located in exon 4 of *MYBPC3*. **(i)** Results from RT-PCR amplification of mRNA. CTRL- is a non-transfected control. **(j,k)** Prediction of RNA splicing alteration and Sanger sequencing result for variant c.492C>T (p.G164G). In panels (b) and (i), we use “n” and “s” to indicate bands corresponding to native splicing, or to skipping of exons, respectively. M corresponds to 1Kb plus DNA ladder (Invitrogen) and base pairs are indicated. See **Supplementary File S2** for experimental details.

In **Figure 2a-g**, we show results from amplification of the exon15-exon21 region using mRNA obtained from blood samples that carry different *MYBPC3* variants. If splicing is correct, amplification results in a fragment of 692 bp, as shown for the wild-type (WT) individual (**Figure 2b**). We observe that mutation c.1624G>C (p.E542Q) leads to a higher-electrophoretic-mobility band at 500 bp, marking skipping of exon 17, in agreement with predictions that native donor site is perturbed by the mutation (**Figure 2c,d,e**). In contrast, the prediction that mutation c.1505G>A (p.R502Q) causes an acceptor site gain was not validated, as splicing proceeded as for the WT (**Figure 2b,f,g,e**). Equivalent results have been obtained before by others (**Table 1**).

In **Figure 2h-k**, we present the results of a mini-gene strategy to test whether the non-pathogenic variant c.492C>T (p.G164G) induces the appearance of a new donor site. RT-PCR amplification shows leads to two bands both in the WT and the mutant sample (**Figure 2i**). The band at 850 bp corresponds to the skipping of the *MYBPC3* insert, a scenario which is not uncommon in mini-gene assays. The band at 1100 bp results from the correct inclusion of exons 4 and 5 of *MYBPC3* in the mini-gene transcript, which was further verified by Sanger sequencing (**Figure 2k**). No other band was amplified in the c.492C>T sample. Hence, the prediction in **Figure 2j** of a new splicing site for variant c.492C>T (p.G164G) was not validated experimentally.

We followed equivalent strategies to examine experimentally all predictions of altered RNA splicing. In total, we were able to collect results for 11 out of the 15 predicted alterations (**Table 1**). For the 4 remaining variants, we could not recruit carriers and the mini-gene assays did not recapitulate native splicing in WT sequences (**Supplementary Figure S3**). RNA splicing alterations leading to premature stop codons could be confirmed experimentally in 6/8 pathogenic mutations (**Figure 1b, Supplementary Table S2**). Validation rate in this set of variants was 5/5 for splicing site losses and 1/6 for splicing site gains. Importantly, none of the predicted alterations of splicing in non-pathogenic variants was confirmed experimentally (**Table 1**). Hence, our results indicate that prediction of RNA splicing alterations and subsequent experimental validation captures at least 14% of the pathogenic mutations with 100% specificity (p = 0.037, Fisher’s exact test, **Figure 1b, Table 1, Supplementary Note S2**).

### Pathogenic mutations induce extensive protein destabilization

A reduction in protein stability leads to more frequent unfolding, which can result in degradation-sensitive polypeptides and reduced total protein levels ^22,38^. To analyze protein destabilization induced by the variants in our database, we used FoldX ^39^. This software can predict protein destabilization if the high-resolution 3D structures of targeted domains are known. Hence, we examined the stability of the variants affecting domains C0, C1, C2, C3, and C5 of cMyBP-C, for which high-resolution structures are available (PDB codes: 2k1m, 3cx2, 1pd6, 2mq0, 1gxe, respectively). We found that the average predicted destabilization induced by pathogenic mutations is higher than for non-pathogenic variants (ΔΔG = 1.8 ± 0.6 *vs* 0.7 ± 0.4 kcal/mol, errors are SEM, p = 0.049, one-sided Student’s t-test). Closer inspection of the data shows that pathogenic mutations p.P187R, p.V471E, p.P645L are predicted to induce extensive reduction of stability, well above the highest destabilization estimated for non-pathogenic variants (**Figure 1d; Supplementary File S1**). The distribution of ΔΔG values for the rest of pathogenic mutants appears indistinguishable from that of non-pathogenic variants (**Figure 1d**).

To validate predictions of extensive domain destabilization induced by pathogenic mutations, we examined experimentally the stability of 18 mutated domains throughout the sequence of cMyBP-C (**Figure 3a**). We analyzed the three pathogenic and non-pathogenic variants with the highest predicted destabilization. We also undertook expression of all variants, regardless of pathogenicity, that target domains for which predictions were not available (**Supplementary Files S1**, see Methods). We did not study at the protein level variants predicted to cause RNA splicing alterations.

**Figure 3.**
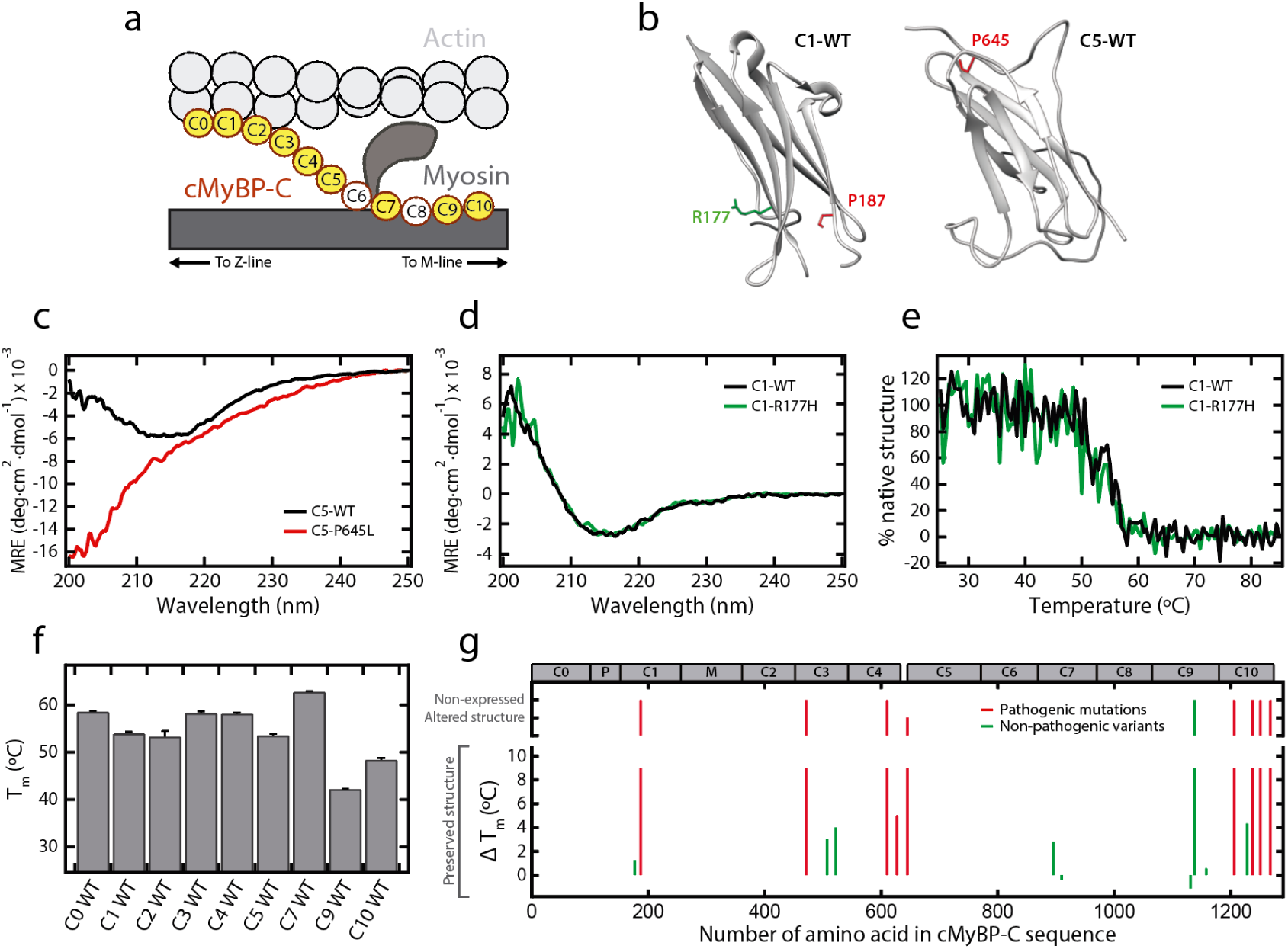
Experimental characterization of protein destabilization induced by *MYBPC3* variants. **(a)** cMyBP-C encompasses 11 domains. To analyze structural destabilization induced by mutations, we produced recombinant versions of WT and mutant domains in *E. coli* (domains not colored could not be expressed). **(b)** Representations of domains C1 (pdb: 3cx2) and C5 (pdb: 1gxe), highlighting positions targeted by 2 pathogenic mutations (in red) and by non-pathogenic variant (in green). Panel was produced with Chimera ^40^. **(c)** CD spectra (given as mean residue ellipticity, MRE) obtained for C5 WT (black), and C5-P645L (red) domains. **(d)** CD spectra obtained for C1 WT (black), and C1-R177H (green) domains. **(e)** Thermal denaturation curves of C1 WT (black), and C1-R177H (green) obtained by tracking the CD signal at 205 nm. The temperature at the midpoint of the transition, T_m_, is obtained from sigmoidal fits to denaturation curves considering a two-state unfolding process. **(f)** T_m_ for WT cMyBP-C domains. **(g)** Change in T_m_ induced by pathogenic (red) and non-pathogenic (green) variants. Plot also indicates mutant domains that could not be expressed or that resulted in altered CD spectrum at 25°C. The raw data for all mutants are presented in **Supplementary Figures S4 and S5**. Position of cMyBP-C domains is indicated at the top of the panel.

The expression of mutated domains was induced in *E. coli* and purified domains were analyzed by far-UV circular dichroism (CD) spectroscopy, a technique that reports on protein secondary structure and stability ^41,42^. We found that 7 out of the 9 pathogenic mutants (C1-P187R, C3-V471E, C4-D610N, C10-G1206D, C10-T1237P, C10-Y1251H, C10-L1268P) could not be produced in soluble form in the most favorable expression conditions, suggesting strong destabilization by the mutations (see Methods, **Figure 3b, Supplementary File S1, Supplementary Figure S4**). Among the pathogenic mutants that could be produced, C5-P645L showed strongly perturbed CD spectrum, indicative of major structural alterations in this mutant (**Figure 3b,c**). The CD spectrum of mutant C4-A627V was more similar to WT, suggesting no major structural alterations in this mutant (**Supplementary Figure S5**). To further characterize the impact of the A627V mutation on domain C4, we tracked the CD signal at increasing temperatures. As the protein domain transitions between the native and the unfolded states, the CD signal varies (**Supplementary Figure S6**) and the temperature at the midpoint of the denaturing transition, or melting temperature (T_m_), informs on the thermal stability of the domain ^42^. We determined that C4-A627V has slightly lower T_m_ than the WT domain (ΔT_m_ = T_m_ (WT) - T_m_ (mutant) = 5.0 °C) (**Supplementary Figure S5**). Regarding non-pathogenic variants, 8 out of preserve protein structure and stability with maximum ΔT_m_ values of 4.3°C (**Figure 3b, d-g, Supplementary Figure S5, Supplementary File S1**). These results indicate that limited changes in T_m_ up to ~5 °C are generally well tolerated and cannot be linked to pathogenicity. There was only one non-pathogenic variant that could not be produced (C9-R1138H) (Supplementary Figure S4). Using molecular dynamics simulations, we obtained evidence that most mutant domains that could not be expressed in native form are indeed destabilized (Supplementary Table S3).

In summary, our workflow to analyze protein stability was able to capture strong domain destabilization in 18% of the pathogenic mutations. Results indicate that domain destabilization can be linked to pathogenicity with 94% specificity (p = 0.048, Fisher’s exact test, **Figures 1b, 3g, Supplementary Note S2**).

### Prevalence of cMyBP-C haploinsufficiency drivers

Our results in the previous sections show that one third of the pathogenic mutations in our database induce alteration of RNA splicing or extensive protein destabilization. The extent of these cMyBP-C molecular phenotypes among pathogenic mutations is probably higher since bioinformatics predictions are not available for all mutants, particularly in the case of protein destabilization (**Supplementary Figure S7**). Hence, we have analyzed the distribution of cMyBP-C haploinsufficiency drivers in the subset of mutants for which we have information for both RNA splicing and protein stability. This analysis shows that 45% of the mutations induce RNA splicing alteration or protein destabilization, and that reduction in protein stability is two times more frequent than splicing defects (**Figure 4**). Interestingly, the majority of mutations do not induce any of these two molecular phenotypes, as observed before for p.R502W ^16,43^, the most common mutant in HCM^13^.

**Figure 4.**
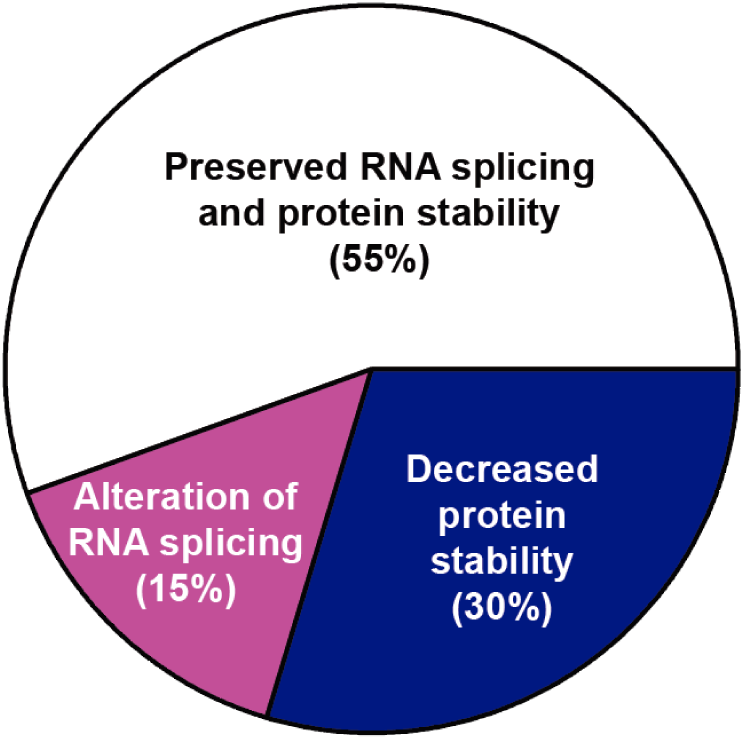
Landscape of molecular phenotypes induced by putative non-truncating pathogenic mutations in *MYBPC3*. For this analysis, we only considered the 27 pathogenic mutations in our database in **Supplementary File S1** for which data on protein stability were available (21 predictions and 6 additional mutations that were studied experimentally in the absence of predictions). Predictions of RNA splicing alteration were available for the 27 pathogenic mutations. All predicted alterations of RNA splicing and protein destabilization were tested experimentally, leading to confirmation of 4 alterations of RNA splicing and 8 highly destabilizing mutations. Our analysis assumes that bioinformatics predictions are able to capture alterations with 100% sensitivity ^7,39^.

### Molecular phenotyping of variants of uncertain significance

Since defects in RNA splicing and protein destabilization are linked to pathogenicity with close to 100% specificity, we investigated whether these molecular phenotypes could be used to inform pathogenicity of putative non-truncating variants currently classified as VUS. To this aim, we studied the 73 VUS with MAF < 10^−4^ reported in ClinVar that target domains C0, C1, C2, C3 and C5 of cMyBP-C (**Figure 5a, Supplementary File S3**). RNA splicing was predicted to be altered in 14/68 variants in which native splice sites were detected, and a new splicing site was predicted in one of the 5 remaining variants (**Figure 5b, Supplementary File S3**). We gathered experimental information for 10 of them, leading to confirmation of protein-damaging splicing alterations in 1/2 predicted splice site losses and in 1/9 predicted site gains, both of them leading to damaging effects at the protein level (**Figure 5c, Table 2, Supplementary Table S2**). Regarding protein stability, 10 variants showed higher predicted destabilization than the most destabilized non-pathogenic variant (**Figure 5d**). We verified experimentally extensive domain destabilization in four of them (C1-G155V, C1-G169S, C1-L199P, and C2-V385M, **Figure 5e, Supplementary Figure S8**). We considered that ΔT_m_ > 10°C marks major impact in protein stability according to previous screenings of destabilization by single amino acid polymorphisms Altogether, we were able to detect alterations of splicing in 2.9% of the VUS for which we had bioinformatics predictions, and up to 3.9% if we consider the expected validation rate of the 5 splicing site gain predictions that could not be tested (**Table 2, Supplementary Note S2**). Protein destabilization was apparent in 5.5% of the variants. Hence, both molecular phenotypes in combination provide strong evidence of pathogenicity of 9% of putative missense and synonymous VUS in *MYBPC3*.

**Table 2.** Experimental validation of predicted alterations of RNA splicing in *MYBPC3* VUS using mini-genes. DL: donor loss; DG: donor gain; AG: acceptor gain. Five variants could not be studied. We studied experimentally the variants for which no information was available in the literature.

| Variant (cDNA) | Variant (Protein) | Predicted splicing alteration | Experimental splicing alteration | Reference |
| --- | --- | --- | --- | --- |
| 39C>T | S13S | DG | - | Supp. File S2 |
| 194C>T | T65M | AG | - | Supp. File S2 |
| 479G>A | R160Q | AG | No | Supp. Figure S2b |
| 505G>A | G169S | DL, DG | No | Supp. Figure S2c |
| 659A>G | Y220C | DG | No | <sup>7</sup> |
| 1213A>G | M405V | DG | Yes | <sup>7,36</sup> |
| 1287G>A | A429A | AG | - | Supp. File S2 |
| 1309G>A | V437M | DG | - | Supp. File S2 |
| 1466A>T | D489V | DG | - | Supp. File S2 |
| 2155T>C | C719R | DG | No | Supp. Figure S2d |
| 2217G>A | E739E | AG | No | Supp. Figure S2d |
| 2234A>G | D745G | DG | No | <sup>7</sup> |
| 2249C>T | T750M | AG | No | Supp. Figure S2d |
| 2288A>G | N763S | DG | No | Supp. Figure S2d |
| 2308G>C | D770H | DL | Yes | Supp. Figure S2d |

**Figure 5.**
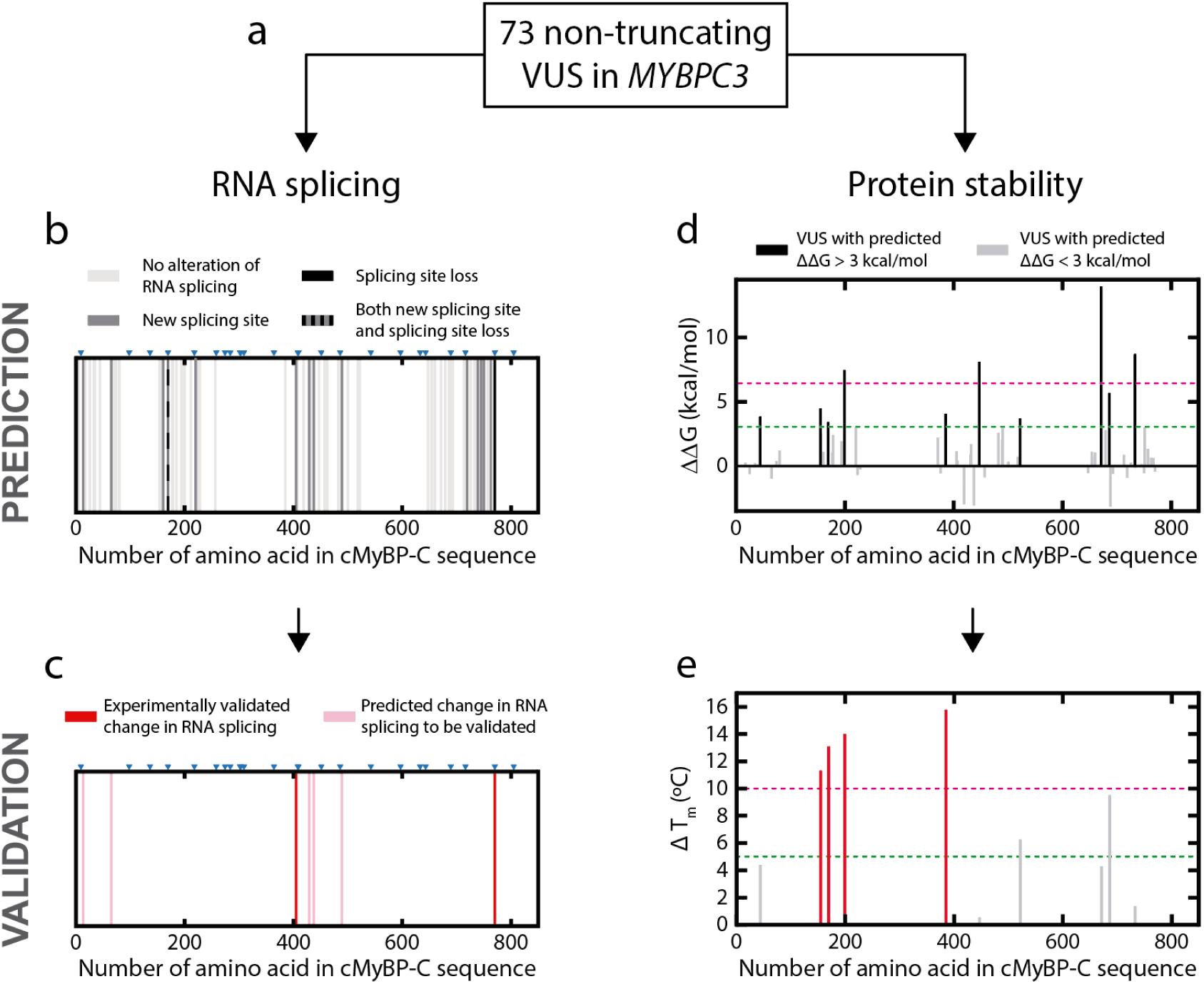
Assessment of cMyBP-C haploinsufficiency drivers in *MYBPC3* VUS. **(a)** 73 VUS in ClinVar were screened for alterations of RNA splicing and protein stability **(b)** Results from predictions of RNA splicing. Blue triangles indicate exon-exon boundaries. Each bar corresponds to a single variant and is colored according to the predicted effect on RNA splicing. **(c)** Experimental validation of predicted changes Variants which could not be tested by mini-gene assays are colored in light pink (see **Table 2**). **(d)** Predicted protein destabilization of the 73 VUS. Each bar corresponds to a single variant and is colored according to the predicted protein destabilization. Reference lines as in Figure 1d. **(e)** Experimental determination of changes in thermal stability for the 10 VUS with predicted ΔΔG > 3 kcal/mol. The green reference line at ΔT_m_ = 5°C marks destabilization values that can be found in non-pathogenic variants (**Figure 3g**), while we consider ΔT_m_ > 10 °C (pink reference line) a signature of pathogenic mutations (corresponding bars are colored in red, while variants below the 10°C threshold are shown in grey). **Supplementary Figure S8** shows the CD data.

## DISCUSSION

The clinical management of HCM has greatly benefited from the discovery of causative genes over the last two decades ^1^. Currently, genetic testing is a Class I recommendation in both the European and American HCM clinical practice guidelines, since identification of pathogenic, actionable genetic variants helps in the clinical care of patients and their families. Thanks to the advent of next-generation DNA sequencing techniques, the number of genetic variants found in HCM patients has increased considerably, leading to new challenges in variant interpretation ^4,33,45–47^. Many genetic variants are present only in a few patients, limiting the power of cosegregation analyses to assess pathogenicity. Alternatively, molecular and functional deficits linked to disease progression can enable classification of rare pathogenic mutations, as already implemented in the ACMG guidelines ^28,45,48–50^.

Here, we have undertaken molecular phenotyping of variants in *MYBPC3* looking for protein haploinsufficiency drivers that can sustain pathogenicity. Importantly, we have compared well-established pathogenic and non-pathogenic variants, which has allowed us to link molecular properties to pathogenicity. Although our strict clinical criteria result in smaller sample sizes than studies analyzing large genetic databases ^7^, our results show that 45% of putative non-truncating pathogenic mutations in *MYBPC3* significantly and specifically induce alteration of RNA processing or extensive protein destabilization (**Figure 4**). Furthermore, we have identified these pathogenicity drivers in *MYBPC3* variants that are currently classified as VUS (**Figure 5**).

Figure 6 summarizes our workflow to assess pathogenicity of *MYBPC3* variants through molecular phenotyping. The first step is to use *in silico* tools to predict alterations in RNA splicing and protein stability. Positive hits are then validated experimentally. Regarding alteration of splicing, we have measured a 14% false positive rate for the prediction of loss of splicing sites, while we have found considerably higher number of false positives for new splicing site prediction, in agreement with previous observations ^7,33^ (**Supplementary Note S2**). Hence, experimental validation of alterations of RNA splicing is a strict requirement to identify true positives, which in the clinical setting an be easily implemented by analyzing the peripheral blood of carriers. We also recommend validation of protein destabilization predictions, since we could only verify extensive protein destabilization in 40% of triaged VUS variants (**Figure 5d,e**). Experimental analysis following negative predictions is discouraged because false negatives are not common in bioinformatics predictions ^7,39^. Direct experimental testing would only be indicated if algorithms fail to capture native splicing sites, or when there is no high-resolution protein structural information. According to our results, the workflow in **Figure 6** can provide strong evidence of pathogenicity for 9% of the *MYBPC3* missense and synonymous variants currently classified as VUS in ClinVar. As a corollary, considering that alteration of RNA splicing and protein thermal stability are found in 45% of pathogenic mutations (**Figure 4**), we can estimate that 20% of missense and synonymous variants in *MYBPC3* currently classified as VUS show strong evidence of pathogenicity.

**Figure 6.**
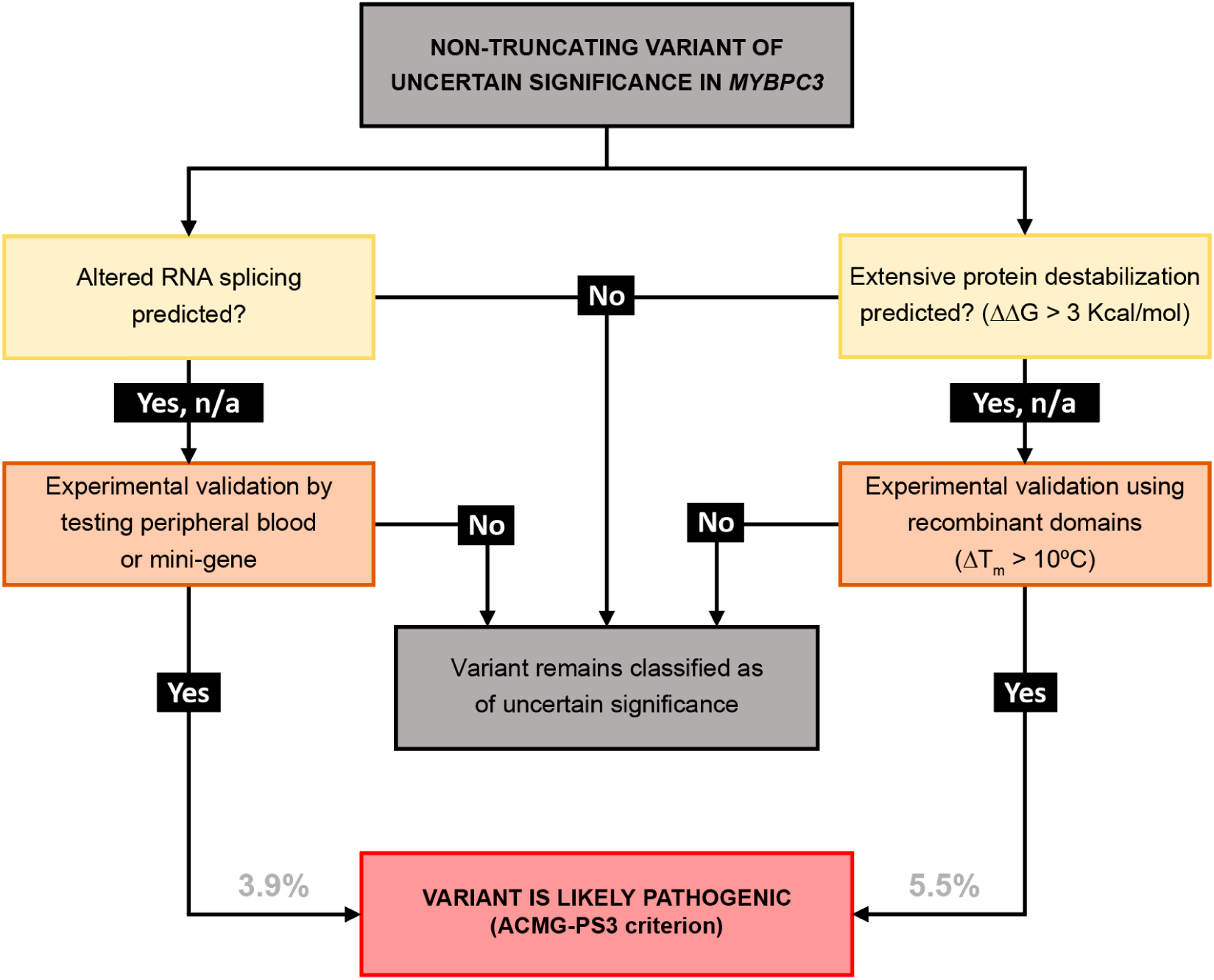
Flow diagram to characterize clinical impact of putative non-truncating VUS in *MYBPC3*. Following bioinformatics predictions of alteration of RNA splicing or protein stability, experimental validation can result in variant reclassification. n/a: predictions not available. Numbers at the bottom indicate the estimated percentage of reclassifications according to our results.

Our study supports the emerging view that genetic variants that affect RNA splicing are pathogenic ^7,8^. Interestingly, these RNA-splicing-altering mutants can be found in exonic and intronic regions that are far from splice sites if the consequence of the mutation is the activation of a new splice site (**Tables 1,2**) ^7,33^. In the clinical setting, evidence of RNA splicing alteration can lead to variant reclassification following ACMG guidelines ^8^. In this context, false positives that misclassify non-pathogenic variants as pathogenic are a source of concern ^5^. Some non-pathogenic variants are found in regions close to splice sites; however, they are not predicted to alter splicing when compared to pathogenic mutations ^8^. Our data agree with this observation. (**Supplementary Note S1**). To the best of our knowledge, there is not a single example of a non-pathogenic variant that has been demonstrated to cause alteration of RNA splicing.

Importantly, we have identified that more than one third of true pathogenic missense *MYBPC3* mutations cause extensive protein destabilization. Remarkably, the number of these variants appears to be higher than the number of exonic mutations affecting RNA splicing (**Figures 4, 5**). However, there are challenges associated with the functional classification of *MYBPC3* variants following protein stability assessment. First, there is no high-resolution structural information for many cMyBP-C domains, which hampers effective triaging of potentially disruptive mutations.

A second challenge stems from the validation of protein destabilizing phenotypes using recombinant domains produced in bacteria, which in the case of very damaging mutations can be inherently impossible. Although our interpretation that strong destabilization hampers domain purification is supported by molecular dynamics simulations (**Supplementary Table S3**), lack of stability is not the only reason why recombinant production of mutant domains may fail ^51^. In our set of experiments, we could not produce the non-pathogenic the C9 domain carrying variant R1138H. The C9 domain has the lowest T_m_ value among all domains assayed (42.2°C, **Figure 3f**). However, C9 is a fibronectin-III type domain, a protein fold whose typical T_m_ values are well above 50°C ^52^. We speculate that our recombinant C9 domain may not recapitulate native stability and that the impact of minor, non-pathogenic changes in stability may be higher in this context. It is also possible that the native environment of the sarcomere limits the destabilizing effect of the mutation via posttranslational modifications or protein-protein interactions. In this regard, stability analysis of cMyBP-C domains that have interaction partners in the sarcomere may be less specific than in the case of central domains of the protein ^53^.

The fact that protein stability is not binary poses a third challenge to protein-destabilization-guided functional classification of variants. We have observed that non-pathogenic variants can cause slight protein destabilization (ΔT_m_ < 5°C, **Figure 3g**). We have considered that ΔT_m_ > 10°C is a signature of pathogenic protein destabilization taking into account the typical distribution of ΔT_m_ in unselected single-amino-acid polymorphisms across different proteins ^44^ The confidence of this threshold could be increased by measuring the stability of more non-pathogenic variants.

Mechanistically, both the alteration of RNA splicing and the destabilization of cMyBP-C domains can lead to reduced cMyBP-C levels, similar to the situation induced by truncating mutations ^15,24,54,55^. It is remarkable though that many synonymous and pathogenic missense mutations in *MYBPC3* do not alter RNA splicing or protein stability (**Figure 4**). The pathogenicity triggers of those mutations remain unknown. A tempting hypothesis is that some of them induce protein haploinsufficiency by alternative mechanisms, including decreased rates of transcription and translation ^56–58^, increased recognition by mRNA and protein degradation machineries ^56,59,60^, and defective incorporation of cMyBP-C in the sarcomere ^61^. Alternatively, mutations can lead to altered binding to protein partners ^62–66^. Tantalizingly, both scenarios could lead to similarly altered modulation of the super relaxed state of myosin, inducing sarcomere hypercontractility typical of HCM ^17,67^.

In summary, we propose that identification of protein haploinsufficiency drivers in *MYBPC3* variants provides strong evidence of their pathogenicity (PS3 criterion in the ACMG guidelines^28^), contributing to the assessment of pathogenicity of putative non-truncating variants of *MYBPC3*. RNA splicing alterations and protein destabilization can be validated using readily available laboratory assays, which addresses the urgent need of methods that assign pathogenicity of genetic variants associated with HCM ^49,68^. Our strategy increases the number of actionable variants in *MYBPC3*, leading to improved clinical management of HCM families.

## MATERIALS AND METHODS

### Human samples

Human samples were obtained following informed consent of patients according to the guidelines of the Declaration of Helsinki. Research involving humans was authorized by the *Comité de Ética de Investigación* of *Instituto de Salud Carlos III* (PI 39_2017).

### Selection of genetic variants

We retrieved *MYBPC3* variants from the *Health in Code (HIC)-Mutations* database, which includes information about >155,000 individuals obtained from >50,000 articles in the literature, as well as from HIC’s own clinical reports. To obtain pathogenicity information from ClinVar, a score was calculated by averaging all available pathogenicity interpretations. With that aim, a scale between 0 and 4 was used for benign (B), likely benign (LB), VUS, likely pathogenic (LP) and pathogenic (P) interpretations. To select VUS in *MYBPC3*, we considered all non-truncating variants associated with hypertrophic cardiomyopathy in ClinVar, excluding those with conflicting interpretations of pathogenicity or showing MAF > 10^−4^. We restricted our analysis to variants targeting domains for which there is high-resolution structural information.

### Bioinformatics prediction of haploinsufficiency drivers

We used Alamut Visual (Interactive Biosoftware) to predict RNA splicing alterations induced by all variants in our pathogenic/non-pathogenic and VUS databases. Alamut implements analyses of four different RNA splicing prediction algorithms (SSF, MaxEnt, NNSPLICE, GeneSplicer). We first determined if at least two of the algorithms identified the canonical splicing site around the mutation site. We then calculated the percent change in splicing score for native sites and for potential new splicing sites. Positive hits result in at least two tools predicting >10% decrease (for native sites) or increase (for new sites) in splicing score, in agreement with published guidelines ^69^. Changes in protein thermodynamic stability were estimated by FoldX. This software estimates changes in free energy (ΔΔG) upon mutation from empirically and statistically derived energy functions^39^.

### Analysis of RNA splicing

For experimental determination of RNA splicing, we resourced to analysis of blood samples from variant carriers or used engineered mini-gene constructs (**Supplementary File S2**). In the case of blood samples, total RNA was extracted from leukocytes of carriers using Trizol (Thermo Fischer Scientific). RNA retro-transcription was done with random primers by SuperScript™ IV VILO™ Master Mix (Thermo Fischer Scientific) and the region of interest was PCR amplified using specific oligos (**Supplementary File S2**). RT-PCR products were purified using the Qiaquick PCR purification Kit (Qiagen) and then Sanger-sequenced when necessary. Correct RNA splicing results in readable electropherograms, obtained for both primers, and whose sequence match the sequence of the canonical cDNA for cMyBP-C. For mini-gene experiments the WT and mutated gDNA fragments were ordered to Integrated DNA Technologies, including at least the exon of interest and the 5′ and 3′ intronic flanking regions (**Supplementary File S2**). The constructs were cloned into β-globin’s intron 2 of pMGene vector^70^ using KpnI. Alternatively, pMGene vectors including the inserts of interest were ordered to GeneArt Gene Synthesis (from Thermo Fisher Scientific). In both cases, resulting constructs were expressed in monolayers of HEK-293 cells cultured in Dulbecco’s modified eagle medium (DMEM, Gibco) supplemented with 10% fetal bovine serum, and 1% penicillin/streptomycin at 37 °C under 5% CO_2_. Cells were transiently transfected with 1 μg of WT or mutated pMGene using FuGENE HD (Promega) according to the manufacturer’s protocol. 24-48 hours after transfection, cells were collected and mRNA was extracted and retrotranscribed, and PCR products were purified and sequenced to compare splicing of WT and mutated constructs as above.

### Protein expression and purification

The cDNAs coding for the cMyBP-C domains and their mutants were cloned from myocardial RNA, produced by PCR-mutagenesis or acquired commercially to Integrated DNA Technologies. Sequences are available in **Supplementary File S4**. cDNAs were cloned in a custom-modified pQE80L expression plasmid (Qiagen) using BamHI and BglII enzymes. Final expression plasmids were verified by Sanger sequencing. Domains were expressed in *E. coli* BLR(DE3). Cultures at OD_600_ = 0.6-1 were induced with isopropyl β-D-1-thiogalactopyranoside (specific conditions in **Supplementary Files S1, S3 and S4**). In general, we found that expression at lower concentrations of IPTG and temperature ≤ 25°C resulted in better yield of purification of challenging-to-express domains, so these conditions were preferred for the expression of mutant domains. Purification of His-tagged domains was achieved by metal affinity and gel filtration chromatographies ^71^. WT domains C6 and C8 were refractory to recombinant expression (data not shown), so variants targeting these domains could not be analyzed. Proteins were eluted from the final size-exclusion chromatography in 20 mM NaPi, pH 6.5 and 63.6 mM NaCl. Proteins were stored at 4°C. Purity of the preparations was evaluated by SDS-PAGE (**Supplementary Figure S4**).

### Circular dichroism

CD spectra were collected using a Jasco J-810 spectropolarimeter. Purified proteins were tested in 20 mM NaPi, pH 6.5 and 63.6 mM NaCl at protein concentrations ranging from 0.1 to 0.5 mg/mL in 0.1-cm-pathlength quartz cuvettes. Protein concentration was obtained from A_280_ values using theoretical extinction coefficients (**Supplementary Files S1, S3, S4**). Spectra were recorded at 50 nm/min scanning speed and a data pitch of 0.2 nm. Four scans were averaged to obtain the final spectra. The contribution of the buffer was subtracted and spectra were normalized by peptide bond concentration. We considered that major changes in the shape of the CD spectrum that cannot be explained by concentration inaccuracies are a signature of strong domain destabilization. To study thermal denaturation, CD signal at a wavelength at which folded and unfolded protein signals were different was monitored as temperature increased from 25 to 85 °C at a rate of 30°C/h (**Supplementary File S4**). Temperature control was achieved using a Peltier thermoelectric system. Changes in CD signal were fit to a sigmoidal function using IGOR Pro (Wavemetrics) to estimate T_m_.

### Molecular Dynamics simulations

Full atom MD simulations of the C1, C3, C4, C5, C9 and C10 cMyBP-C domains (WT and their variants, see **Supplementary Table S3**) in dodecahedral boxes (1 nm minimal distance between protein atoms and box edges) filled with Tip3p water molecules were performed at 410 K in protonation conditions mimicking pH 7.0 using the Charmm27+CMAP force field as described ^72^. In the absence of high-resolution structures, we resourced to homology models obtained using SwissModel ^73^. Mutations were modelled on the WT structures using SwissPdbViewer v4.1.0 ^73^. For each WT and variant structure, three 1-μs long trajectories were obtained. Trajectories were analyzed using the model described in ^74^ to obtain estimations of ΔΔG upon mutation at 298 K.

**Supplementary Information** is available in the online version of the article. Supplementary info includes 4 independent Supplementary Files, 8 Supplementary Figures, 2 Supplementary Notes, 3 Supplementary Tables and Supplementary References.

## Data Availability

Data is available on reasonable request to the corresponding author

## Acknowledgements

JAC acknowledges funding from the *Ministerio de Ciencia e Innovación* (MCIN) through grants BI02014-54768-P, BI02017-83640-P (AEI/FEDER UE), EIN2019-102966 and RYC-2014-16604, the European Research Area Network on Cardiovascular Diseases (ERA-CVD/ISCIII, MINOTAUR, AC 16/00045) and the *Comunidad de Madrid* (consortium Tec4Bio-CM, S2018/NMT-4443, FEDER). The CNIC is supported by the *Instituto de Salud Carlos III* (ISCIII), MCIN and the Pro CNIC Foundation, and is a Severo Ochoa Center of Excellence (SEV-2015-0505). We acknowledge funding from ISCIII to the *Centro de Investigación Biomédica en Red* (CIBERCV), CB16/11/00425. LS acknowledges funding from MCIN (BFU2015-63571-P). JS acknowledges funding from MCIN (BFU2016-78232-P), Gobierno de Aragón (E45_17R) and ERDF-InterregV-A POCTEFA (PIREPRED-EFA086/15). CSC is the recipient of an FPI-SO predoctoral fellowship BES-2016-076638. MRP was the recipient of a PhD fellowship from the Italian Ministry of Education, Universities and Research (MIUR). HGC is recipient of the FPU16/04232 doctoral contract from MCIN. We thank Natalia Vicente for excellent technical support (through grant PEJ16/MED/TL-1593 from *Consejería de Educación, Juventud y Deporte de la Comunidad de Madrid* and the European Social Fund). We thank the Spectroscopy and Nuclear Magnetic Resonance Core Unit at CNIO for access to CD instrumentation. We thank Andrea Thompson and Sharlene Day for critical reading of the manuscript. We thank all members of the *Molecular Mechanics of the Cardiovascular System* team for helpful discussions and four anonymous reviewers for constructive feedback.

## Author contributions

JAC conceived the project. DGG, PGP, MRP, JAC and LM classified genetic variants according to pathogenicity. MRP and JAC did bioinformatics analysis of RNA splicing. JD, JAC and LS predicted changes in protein stability. DSO, SV, FD, JRG, MSM, RBV, GF, PGP and JAC ensured procurement of human samples. MRP and CSC did experimental analysis of RNA splicing. CSC and DVC cloned and purified proteins. CSC and EHG did circular dichroism experiments. JJGF, HGC and JS did molecular dynamics simulations. CSC, MRP and JAC drafted the manuscript with input from all authors.

## Competing financial interests

LM is share-holder of Health in Code.

## Notes

### Funding Statement

JAC acknowledges funding from the Ministerio de Ciencia e Innovación (MCIN) through grants BIO2014-54768-P, BIO2017-83640-P (AEI/FEDER, UE), EIN2019-102966 and RYC-2014-16604, the European Research Area Network on Cardiovascular Diseases (ERA-CVD/ISCIII, MINOTAUR, AC16/00045) and the Comunidad de Madrid (consortium Tec4Bio-CM, S2018/NMT-4443, FEDER). The CNIC is supported by the Instituto de Salud Carlos III (ISCIII), MCIN and the Pro CNIC Foundation, and is a Severo Ochoa Center of Excellence (SEV-2015-0505). We acknowledge funding from ISCIII to the Centro de Investigación Biomédica en Red (CIBERCV), CB16/11/00425. LS acknowledges funding from MCIN (BFU2015-63571-P). JS acknowledges funding from MCIN (BFU2016-78232-P), Gobierno de Aragón (E45_17R) and ERDF-InterregV-A POCTEFA (PIREPRED-EFA086/15). CSC is the recipient of an FPI-SO predoctoral fellowship BES-2016-076638. MRP was the recipient of PhD fellowship from the Italian Ministry of Education, Universities and Research (MIUR). HGC is recipient of the FPU16/04232 doctoral contract from MCIN.

